# Differences in cardiac vagal modulation and cortisol response in adolescents with and without Autism Spectrum Disorder

**DOI:** 10.1101/2022.09.28.22280456

**Authors:** Anoushka Thoen, Kaat Alaerts, Jean Steyaert, Sophie Pleysier, Tine Van Damme

## Abstract

**Purpose:** Previous research pointed towards a need of enhanced understanding of cardiac vagal modulation during resting and stress conditions in individuals with Autism Spectrum Disorder (ASD). This cross-sectional study addressed the following hypotheses: lower values of cardiac vagal modulation will be found in adolescents with ASD in comparison to typically developing (TD) peers; different levels of cardiac vagal reactivity and recovery will be found in adolescents with ASD; lower cardiac vagal modulation in adolescents with ASD is associated with lower psychosocial functioning and higher cortisol levels.

**Methods:** Age and gender matched groups of adolescents (13-17 year) with ASD (n=47) and TD peers (n=47) were included. Heart rate, breathing frequency and cortisol levels were determined during baseline and a standardized stress-provoking assessment. Behavioral data concerning autism and behavioral characteristics were collected prior to the assessment.

**Results:** Adolescents with ASD displayed lower levels of cardiac vagal modulation during baseline and stress-provocation compared to their TD peers. However, levels of cardiac vagal reactivity and recovery were similar across groups. Weak to moderate associations were found between the level of cardiac vagal modulation and self- and parent-reported measures of autism characteristics and psychosocial functioning in adolescents with ASD. No significant associations were found between baseline cortisol levels and cardiac vagal modulation in both groups.

**Conclusion:** These findings suggest a parasympathetic hypo-activity in adolescents with ASD and, although the level of reactivity and recovery was the same as TD peers, this hypo-activity is related to several aspects of psychosocial functioning.

---

Alterations in the functioning and coordination of the stress system may be, at least partially, related to the core characteristics of Autism Spectrum Disorder (ASD) and the presence of co-occurring conditions; including anxiety and depression as demonstrated in previous research (Hollocks et al., 2014; Muscatello et al., 2021). The stress system contains two major interconnected peripheral components, namely the autonomic nervous system (ANS) and the hypothalamic-pituitary-adrenal (HPA) axis (Agorastos et al., 2018; Thayer &Lane, 2000). The ANS is composed of two branches, the parasympathetic and the sympathetic nervous system. Traditionally, the parasympathetic nervous system is associated with growth and restorative functions (‘rest and digest’), whereas the sympathetic nervous system is involved in promoting increased metabolic output to deal with challenges from outside of the body (‘fight and flight’; Lydon et al., 2016). Despite their antagonistic function, both subsystems are constantly active and work in a dynamic balance to regulate multiple visceral mechanisms among other bodily functions (Bujnakova et al., 2016). Heart rate variability and respiratory sinus arrhythmia (RSA) are considered important parameters of ANS functioning. Heart rate variability refers to the natural variation between heartbeats and is under the control of both branches of the ANS. RSA is a component of heart rate variability and provides a sensitive index of cardiac vagal (parasympathetic) modulation (Grossman et al., 1990; Lehrer et al., 2020). Cardiac vagal modulation has been linked to positive social functioning, better emotional processing, and better behavioral regulation in both clinical and non-clinical populations (Beauchaine, 2015; Benevides &Lane, 2015; Condy et al., 2019; Edmiston et al., 2016; Makris et al., 2022; Morrish, 2019; Neuhaus et al., 2016; Patriquin et al., 2014; Patriquin et al., 2015; Patriquin et al., 2011). These findings have been based on two theoretical constructs, namely Porges’ Polyvagal Theory (Porges, 1995) and the Neurovisceral Integration Theory (Friedman, 2007; Thayer &Lane, 2009). In short, Porges’ Polyvagal Theory discusses the influence of the myelinated vagal nerve on social behavior within the social engagement system, referring to a collection of muscles necessary for appropriate social interactions (Porges, 2011). This theory proposes that differences in the social engagement system of individuals with ASD coincide with reduced cardiac vagal modulation (Arora et al., 2021). The Neurovisceral Integration Theory describes a Central Activation Network (CAN) consisting of cortical structures (such as prefrontal cortex, anterior cingulate cortex, amygdala and insula) which are related to different aspects of self-regulatory behavior and emotional regulation (Fiskum, 2019). Structural, functional and connectivity differences have been found in individuals with ASD in all these cortical structures, leading to an increased relevance of the Neurovisceral Integration Theory in autism research (Kushki et al., 2014).

Prior studies examining the functioning of the parasympathetic nervous system in children and adolescents with ASD mostly converged on the identification of lower values of RSA during baseline measures as well as during stress conditions, compared to typically developing (TD) peers (Cheng et al., 2020; Edmiston et al., 2016; Lory et al., 2020; Makris et al., 2022; Thapa et al., 2019; Tonhajzerova et al., 2021). However, less conclusive evidence is available for differences in task-based phasic cardiac vagal modulation (referred to as reactivity and recovery levels) in children and adolescents with and without ASD (Benevides &Lane, 2015). Some studies reported no differences in cardiac vagal reactivity and recovery levels between both groups during social and non-social stress-provoking tasks while the overall levels of cardiac vagal modulation were lower in children and adolescents with ASD (Edmiston et al., 2017; Levine et al., 2012; Lory et al., 2020). However, the study of Muscatello et al. (2021) reported an age-effect on cardiac vagal reactivity levels with only older adolescents with ASD showing a blunted response towards a social stress-provoking task in comparison with TD peers. In contrast, lower reactivity levels of cardiac vagal modulation during social stress-provoking tasks have also been reported in children and adolescents with ASD irrespective of the overall level of cardiac vagal modulation (Corbett et al., 2019; Van Hecke et al., 2009). In the majority of studies regarding children and adolescents with ASD, the authors concluded that the extent and direction of change in cardiac vagal modulation during stressors may be different in comparison with TD peers (Benevides &Lane, 2015). However, the use of different outcome measures, the choice of different stress-provoking tasks and the level of emotional relevance and attention given to or required for the tasks make it difficult to compare the studies (Beauchaine et al., 2019; Benevides &Lane, 2015). Furthermore, supporting Porges’ Polyvagal Theory and the Neurovisceral Integration theory, parasympathetic hypo-activity in individuals with ASD has also been associated with the presence of internalizing symptoms (Arora et al., 2021), lower social-emotional skills (Arora et al., 2021; Makris et al., 2022; Patriquin et al., 2019), the presence of more severe auditory and visual hyperreactivity (Arora et al., 2021) and the presence of more repetitive and restrictive behaviors (Condy et al., 2019).

The second major component of the stress system is the HPA axis, which is a neuroendocrine system. It includes a complex set of influences and interactions that result in the secretion of the glucocorticoid cortisol (Agorastos et al., 2018). Cortisol influences all major homeostatic systems of the body, thus resulting in changes in cardiovascular function, immunity, metabolism, and neurobiology (Chrousos, 2009; Sapolsky et al., 2000). Regarding children and adolescents with ASD, a higher between- and within-subject variability of basal cortisol levels have been found as opposed to cortisol levels in TD peers (Taylor &Corbett, 2014). Furthermore, differences in HPA axis responsivity to psychosocial stressors have been commonly reported in individuals with ASD compared to TD peers but no conclusive evidence could be found for its direction (hypo-or hyperactivity; Makris et al., 2022).

In sum, an enhanced understanding of cardiac vagal modulation and HPA axis activity during resting and stress conditions as well as cardiac vagal reactivity and recovery levels in individuals with ASD is needed. The most important methodological shortcomings in previous research are differences in outcome measures; use of small sample sizes; not taking into account the use of medications; differences in severity of autism characteristics and the presence of co-occurring disorders which hinder the comparison of different studies (Arora et al., 2021; Makris et al., 2022). Multiple researchers have therefore recommended to take these confounding factors into account to elucidate findings on cardiac vagal modulation, the HPA axis and the influence on severity of autism characteristics and psychosocial functioning in individuals with ASD (Arora et al., 2021; Benevides &Lane, 2015; Condy et al., 2019; Corbett et al., 2021; Makris et al., 2022; Patriquin et al., 2019).

The main outcome measure of this cross-sectional study is cardiac vagal modulation, a parameter of parasympathetic functioning. Since adolescence is considered as a particularly vulnerable period in terms of stress and current literature in this developmental period is scarce; this specific age group was addressed. Based on previous research findings (Arora et al., 2021; Benevides &Lane, 2015; Cheng et al., 2020; Condy et al., 2019; Makris et al., 2022; Patriquin et al., 2019), three hypotheses were formulated: (1) lower values of cardiac vagal modulation will be found in adolescents with ASD during baseline measurements and two stress-provoking tasks in comparison to TD peers; (2) different levels of cardiac vagal reactivity and recovery will be found in adolescents with ASD in comparison to TD peers; (3) lower values of cardiac vagal modulation as well as reactivity and recovery levels in individuals with ASD are associated with psychosocial functioning (e.g. social difficulties, repetitive behaviors and anxiety) and higher baseline cortisol levels.

## Methods

Data was collected as part of a larger randomized controlled trial as described in Thoen et al. (2021). The current study included adolescents with ASD and TD peers, who were recruited from December 2019 until April 2022.

### Participants

An a priori power analyses (α=0.05; 1-β=0.80) based on a medium effect size (*g*=0.59) as reported in Cheng et al. (2020) was used, recommending at least 37 adolescents (12-18 years) per group. A formal diagnosis of ASD was based on the criteria of the diagnostic and statistical manual of mental disorders (DSM-IV/5; American Psychiatric Association, 2000, 2013). The use of medication and the presence of co-occurring disorders was registered elaborately (see Results section). Exclusion criteria for both groups were (i) the presence of an intellectual disability as described in the DSM-IV/5; (ii) the presence of an uncorrected hearing- or vision impairment; (iii) the presence of congenital heart diseases, diagnosed cardiovascular abnormalities or somatic diseases with a known impact on heart function; (iv) pregnancy; (v) insufficient knowledge of the Dutch language and (vi) participation in other clinical trials. The Autism Quotient – Adolescent Version (Baron-Cohen et al., 2006) was used as an exclusion tool for the TD group in addition to the presence of a neurodevelopmental disorder or psychiatric disorder as described in the DSM-IV/5. Furthermore, TD adolescents with a sibling diagnosed with a neurodevelopmental disorder as described in the DSM-IV/5 were excluded as well.

Recruitment of adolescents with ASD across Flanders (Belgium) was facilitated by the Leuven Autism Expertise Centre and the Leuven Autism Research Group (KU Leuven). In addition, (special education) schools, autism advocacy organizations and support groups as well as independent clinical practices were addressed. TD adolescents were recruited in the same region by contacting local schools and through public advertisement.

### Procedure

Informed parental consent and adolescent assent was obtained from all participants. All questionnaires were filled out online prior to the assessment, except for the visual analogue scale on perceived stress that was administered after the assessment (see ‘Materials’ section).

#### Assessment

The assessment of one hour included a preparatory phase to attach the sensors for physiological data-collection, a stress-provoking protocol, and the collection of saliva samples. A standardized stress-provoking protocol (30 minutes) was used during which heart rate and breathing frequency were continuously registered. To rule out the influence of the cortisol awakening response, the assessments were performed in the afternoon (Hollocks et al., 2014). A detailed description of the assessment (f.i. standardized body positioning and outcome measures) is described in Thoen et al. (2021).

The first ten minutes of the stress-provoking protocol refer to the baseline period during which the adolescents had to sit quietly with their eyes closed. Of note, following the recommendations of Quintana et al. (2016), the first five minutes were excluded from data-analyses to incorporate a corresponding acclimatization period. Subsequently, the ‘Stroop Word-Color Interference task’ (Kushki et al., 2013; Stroop, 1935) and the ‘Social Stress Recall Task’ (SSRT; Bishop-Fitzpatrick et al., 2017; Richman et al., 2007) were used as stress-provoking tasks, each followed by a 5-minute resting period. Both tasks had a duration of five minutes and had been used previously to provoke stress in individuals with ASD (Bishop-Fitzpatrick et al., 2017; Kushki et al., 2013).

### Materials

#### Apparatus and physiological measurement

All physiological data for this study were captured using the NeXus-10 MKII biofeedback device and Biotrace+ Software (Mind Media B.V., The Netherlands). Nexus’ proprietary accessories were used to register heart rate and breathing frequency. Three-lead ECG data was collected with disposable, self-adhesive and pre-gelled electrodes (Kendall™ ECG Electrodes Arbo™ H124SG, Covidien, Ireland) without specific skin preparations at a sampling rate of 256 SPS. The negative electrode was placed medial to the right coracoid process, just below the clavicle. The neutral electrode was placed similarly on the contralateral side. The positive electrode was placed at the lower left ribcage. An elastic band with stretch-sensitive sensors around the waist was used for the recording of the breathing frequency at a sampling rate of 32 SPS.

#### Cortisol

Salivette® Cortisol cotton swabs (Sarstedt Inc., Rommelsdorf, Germany) were used for the collection of saliva samples at three time points to determine the level of cortisol. The baseline sample was collected before the start of the stress-provoking protocol. Since salivatory cortisol levels peak at around 10-30 minutes after stress cessation (Foley &Kirschbaum, 2010), the second and third saliva samples were collected at 20 minutes after each of the stress-provoking tasks (Corbett et al., 2019). Afterwards, the samples were stored under appropriate controlled conditions (−20°C).

#### Psychosocial functioning and self-perceived stress

The parent-reported Social Responsiveness Scale (SRS-2; Constantino &Gruber, 2015; Constantino &Gruber, 2012), Repetitive Behavior Scale (RBS-R Dutch translation; Lam &Aman, 2007) and Autism Quotient (AQ Dutch translation; Baron-Cohen et al., 2006) were used to evaluate autism characteristics. The total score of the latter was also used as an exclusion criterion for adolescents from the TD group in case they exceeded the clinical cut-off score of 32. Furthermore, psychosocial problems and strengths were assessed using the parent- and self-reported Strengths and Difficulties Questionnaire (SDQ Dutch translation; Goodman, 2001). Self-report tools were filled out by the adolescents to report on daily perceived stress levels (Perceived Stress Scale – Adolescent version; Van der Ploeg, 2013), the presence of negative emotions (Depression, Anxiety and Stress Scale – 21; de Beurs et al., 2001) and the level of physical activity during the previous week (Physical Activity Vital Sign – Dutch translation; Greenwood et al., 2010). An indication on the level of sensory hypersensitivity was assessed using a visual analogue scale going from zero (no sensory hypersensitivity) to ten (severe sensory hyperresponsivity and negative impact on functioning). The level of perceived stress for each of the stress-provoking tasks was also evaluated using a visual analogue scale ranging from zero (no stress) to ten (highest level of perceived stress possible).

### Data preprocessing

Heart rate data were preprocessed using Kubios HRV Premium (version 3.4.3, University of Eastern Finland, Kuopio, Finland) using the automatic artifact correction algorithm and medium automatic noise detection. In addition, visual inspection of the raw data was performed. Analysis of the data regarding heart rate variability was also performed with Kubios HRV Premium using respiratory sinus arrhythmia as a measure of cardiac vagal modulation. This was calculated using the time domain measure ‘Root Mean Square of Successive Differences between normal heartbeats’ (RMSSD) and the frequency domain measure ‘High Frequency Heart Rate Variability’ (HF-HRV; Laborde et al., 2017; Malik, 1996). Researcher-developed scripts in MATLAB R2020b (MathWorks, Natick, Massachusetts, USA) were used for preprocessing and calculation of the mean respiratory frequency for the five separate periods in the stress-provoking protocol (Baseline, Stroop, Rest1, SSRT and Rest2).

### Data analyses

Statistical analyses were conducted using SPSS software (IBM SPSS Statistics, version 27). Physiological and cortisol data were logarithm-transformed where needed to meet normality assumptions prior to data analyses. A repeated-measures ANOVA was used with group (ASD and TD) as a between-subjects factor and phase (Baseline, Stroop, SSRT, Rest1 and Rest2) as a within-subjects factor for the analyses of physiological and cortisol data. In cases of violation of sphericity, degrees of freedom were Greenhouse-Geisser or Huynh-Feldt corrected (ε<.75 and ε>.75 respectively). Partial η^2^ was calculated to measure the effect size. Bonferroni-corrections were applied for post-hoc analyses. The level of RSA reactivity and recovery (i.e., difference between a resting period and a stress-provoking task and vice versa, respectively) was calculated and correlated with baseline RSA using Pearson or Spearman’s correlations (Laborde et al., 2017). Finally, group-based exploratory Pearson and Spearman’s correlations were used to examine the relationship between RSA (at baseline, reactivity and recovery levels) and measures of psychosocial functioning and the baseline cortisol level.

A control-analysis with another measure reflecting cardiac vagal tone, HF-HRV (range between 0.15 and 0.4 Hz as calculated in Kubios HRV), was performed using independent sample t-Tests and Mann-Whitney U-Tests for Baseline, Rest1 and Rest2 with Cohen’s *d* and r, respectively as measures of effect size. This control-analysis follows the recommendations of Laborde et al. (2017) and Malik (1996) to ensure that the results echo the findings of RMSSD. Since HF-HRV should not be used as a measure of cardiac vagal activity in tasks where verbal responses are required (Thomas et al., 2019), the data of the Stroop and SSRT were not included in these analyses. Missing values were excluded from the analyses.

## Results

In total, 47 adolescents with ASD and 47 gender and age matched TD peers were included in the analyses. Demographic and diagnostic information is presented in Table 1. [Please insert Table 1 here] Independent samples t-tests revealed significant group differences on the AQ, SRS-2, RBS-R, SDQ (parent- and self-report), DASS-21, PSS and the visual analogue scale for sensory hypersensitivity (*p*>0.001). In both groups, use of contraceptive medication (TD n=8; ASD n=4), sleep medication (TD n=1; ASD n=2) and medication related to allergies, somatic or neurological problems (TD n=3; ASD n=7) was reported. Only adolescents with ASD reported the use of psychotropic medication: methylphenidate (n=7), non-tricyclic antidepressants (n=8), antipsychotic medication (n=11) and anti-epileptic medication (n=3). Co-occurring neurodevelopmental and/or psychiatric disorders were present in 22 adolescents with ASD, namely: attention deficit (hyperactivity) disorder (n=12), obsessive compulsive disorder (n=4), bipolar disorder (n=1), depression (n=2), anxiety disorder (n=4), eating disorder (n=1), trauma (n=1) and developmental coordination disorder (n=4).

**Table 1.** Participant demographics by group

|  | ASD (n = 47; 25 females) |  |  | TD (n = 47; 25 females) |  |  | *p* | Cohen's *d* |
| --- | --- | --- | --- | --- | --- | --- | --- | --- |
|  | M | SD | Range | M | SD | Range |  |  |
| Age (years) | 15.88 | 1.79 | 12.5-18.8 | 15.89 | 1.80 | 12.6-18.9 | 0.958 | 0.011 |
| AQ total score | 31.65<sup>a</sup> | 7.39 | 10-48 | 9.47 | 5.38 | 1-23 | <0.001 | 3.438 |
| SRS-2 total score (t-score) | 85.77 | 15.78 | 52-111 | 47.32 | 5.94 | 34-65 | <0.001 | 3.225 |
| RBS-R total score | 15.98 | 11.95 | 1-52 | 1.53 | 2.09 | 0-8 | <0.001 | 1.684 |
| SDQ total score parent-report | 15.54<sup>a</sup> | 6.18 | 4-30 | 4.40 | 3.25 | 0-16 | <0.001 | 2.264 |
| SDQ total score self-report | 16.72 | 5.76 | 4-27 | 8.72 | 4.67 | 0-19 | <0.001 | 1.525 |
| DASS-21 | 55.02 | 29.31 | 2-120 | 25.15 | 17.97 | 0-70 | <0.001 | 1.229 |
| PSS-Adolescent version | 24.62 | 6.38 | 8-38 | 17.23 | 5.08 | 6-28 | <0.001 | 1.280 |
| Sensory hypersensitivity | 6.13 | 2.63 | 0-10 | 3.04 | 2.54 | 0-7 | <0.001 | 1.193 |
| PAVS (total minutes per week) | 165.14<sup>b</sup> | 132.37 | 0-525 | 220.29<sup>c</sup> | 133.59 | 0-600 | 0.085 | 0.415 |

Missing value: <sup>a</sup>(n=1); <sup>b</sup>(n=10); <sup>c</sup>(n=13). ASD=Autism Spectrum Disorder; AQ=Autism Quotient; DASS=Depression Anxiety and Stress Scale; M=Mean; PAVS=Physical Activity Vital Sign; PSS=Perceived Stress Scale; RBS-R=Repetitive Behavior Scale-Revised; SD=Standard Deviation; SDQ=Strengths and Difficulties Questionnaire; SRS-2=Social Responsiveness Scale-Second Edition; TD=Typically Developing

Descriptive statistics (mean and standard deviation) for heart rate, cardiac vagal modulation, breathing frequency and cortisol levels are presented per group in Table 2. [Please insert Table 2 here]

**Table 2.**
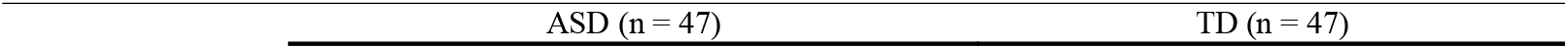
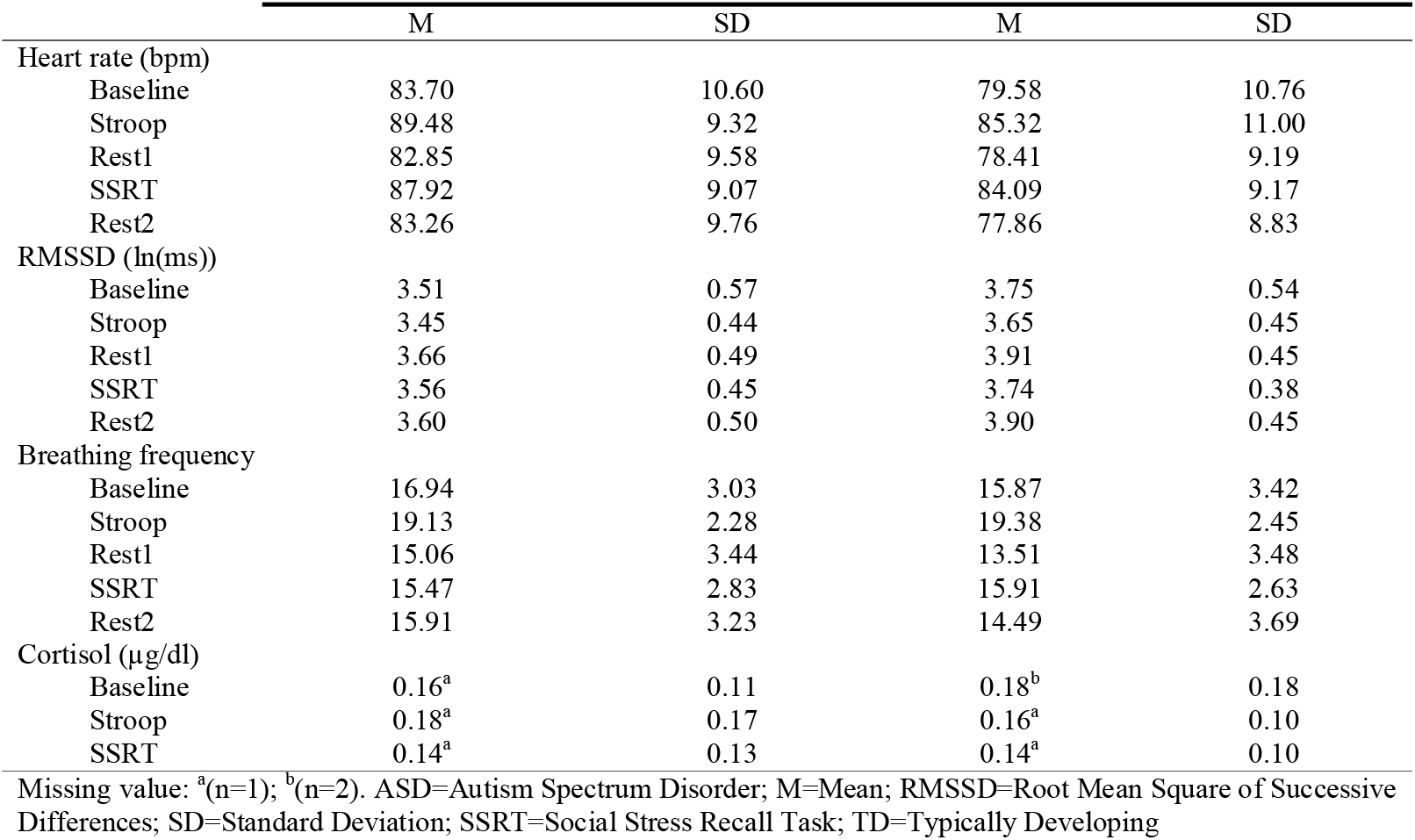
Mean and Standard Deviation of heart rate, cardiac vagal modulation, breathing frequency and cortisol levels per group

|  | M | SD | M | SD |
| --- | --- | --- | --- | --- |
| Heart rate (bpm) |  |  |  |  |
| Baseline | 83.70 | 10.60 | 79.58 | 10.76 |
| Stroop | 89.48 | 9.32 | 85.32 | 11.00 |
| Rest1 | 82.85 | 9.58 | 78.41 | 9.19 |
| SSRT | 87.92 | 9.07 | 84.09 | 9.17 |
| Rest2 | 83.26 | 9.76 | 77.86 | 8.83 |
| RMSSD (ln(ms)) |  |  |  |  |
| Baseline | 3.51 | 0.57 | 3.75 | 0.54 |
| Stroop | 3.45 | 0.44 | 3.65 | 0.45 |
| Rest1 | 3.66 | 0.49 | 3.91 | 0.45 |
| SSRT | 3.56 | 0.45 | 3.74 | 0.38 |
| Rest2 | 3.60 | 0.50 | 3.90 | 0.45 |
| Breathing frequency |  |  |  |  |
| Baseline | 16.94 | 3.03 | 15.87 | 3.42 |
| Stroop | 19.13 | 2.28 | 19.38 | 2.45 |
| Rest1 | 15.06 | 3.44 | 13.51 | 3.48 |
| SSRT | 15.47 | 2.83 | 15.91 | 2.63 |
| Rest2 | 15.91 | 3.23 | 14.49 | 3.69 |
| Cortisol (μg/dl) | | | | |
| Baseline | 0.16<sup>a</sup> | 0.11 | 0.18<sup>b</sup> | 0.18 |
| Stroop | 0.18<sup>a</sup> | 0.17 | 0.16<sup>a</sup> | 0.10 |
| SSRT | 0.14<sup>a</sup> | 0.13 | 0.14<sup>a</sup> | 0.10 |

Missing value: <sup>a</sup>(n=1); <sup>b</sup>(n=2). ASD=Autism Spectrum Disorder; M=Mean; RMSSD=Root Mean Square of Successive Differences; SD=Standard Deviation; SSRT=Social Stress Recall Task; TD=Typically Developing

### Cardiac vagal modulation: adolescents with ASD and TD peers

Repeated measures ANOVA with group as a between-group variable and phase as a within-group variable revealed a significant effect on cardiac vagal modulation (expressed as RMSSD) over time (F(2.625,241.461)=17.058, *p*<.001, η^2^_p_=.156) but no significant phase x group interaction effect (*p*=.371). However, a main effect of group was identified (F(1,92)=7.020, *p*=.009, η^2^_p_=.071), indicating that irrespective of the phase, adolescents with ASD presented significantly lower cardiac vagal modulation with respect to their TD peers (see Figure 1). [Please insert Fig.1 here]

**Fig. 1.**
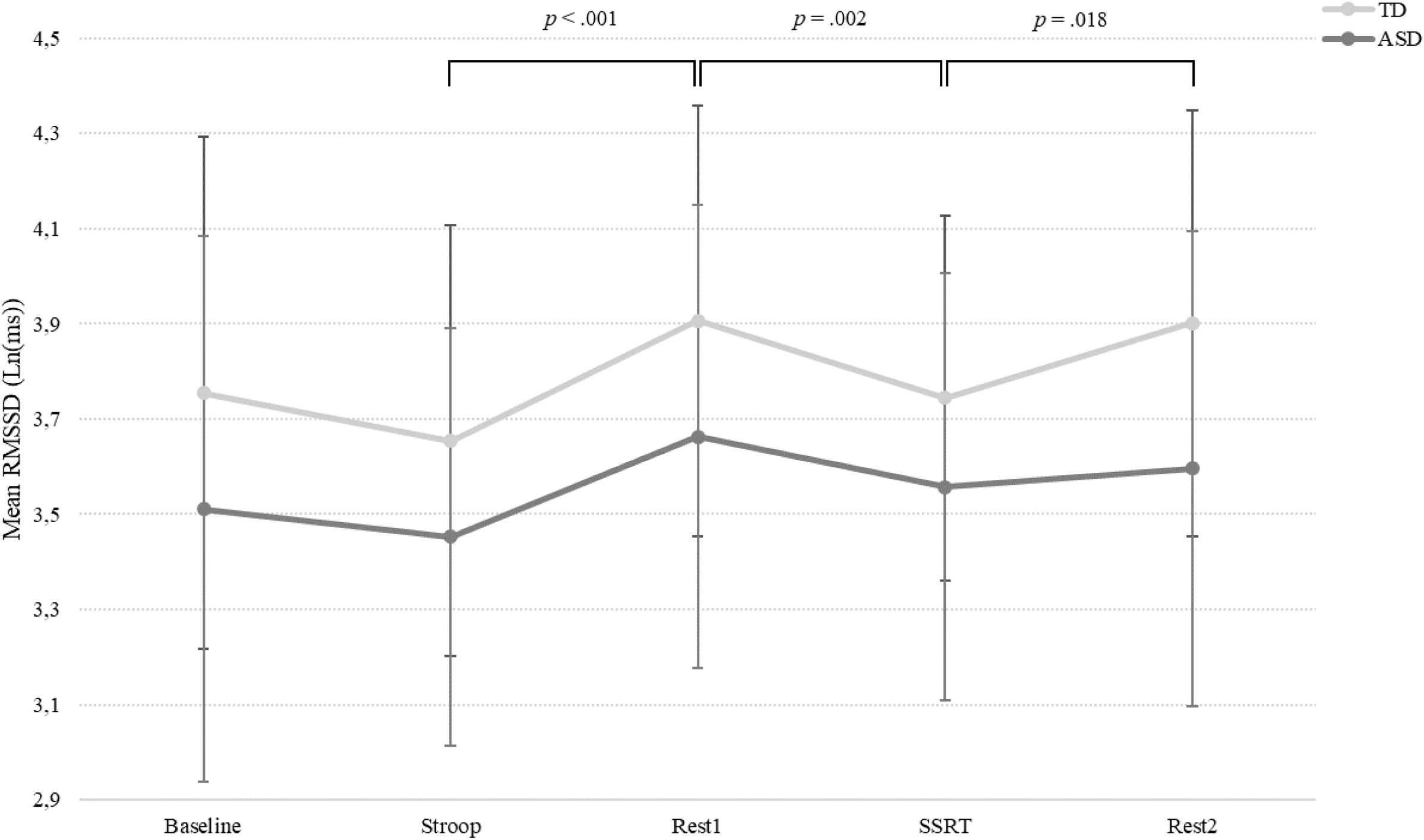
Change in cardiac vagal modulation, expressed as RMSSD, during the stress-provoking assessment. Note: ASD=Autism Spectrum Disorder, RMSSD=Root Mean Square of Successive Differences; SSRT=Social Stress Recall Task, TD=Typically Developing

Weak to strong correlations were found between baseline cardiac vagal modulation and reactivity and recovery levels in both groups (*p*<.01; see Table 3). [Please insert Table 3 here] Higher baseline values were associated with larger reductions of cardiac vagal modulation during stress-provoking tasks and larger increases of this parameter during recovery (visualizations are available as Supplementary Material). [Please provide electronically access towards Supplementary Material]

**Table 3.**
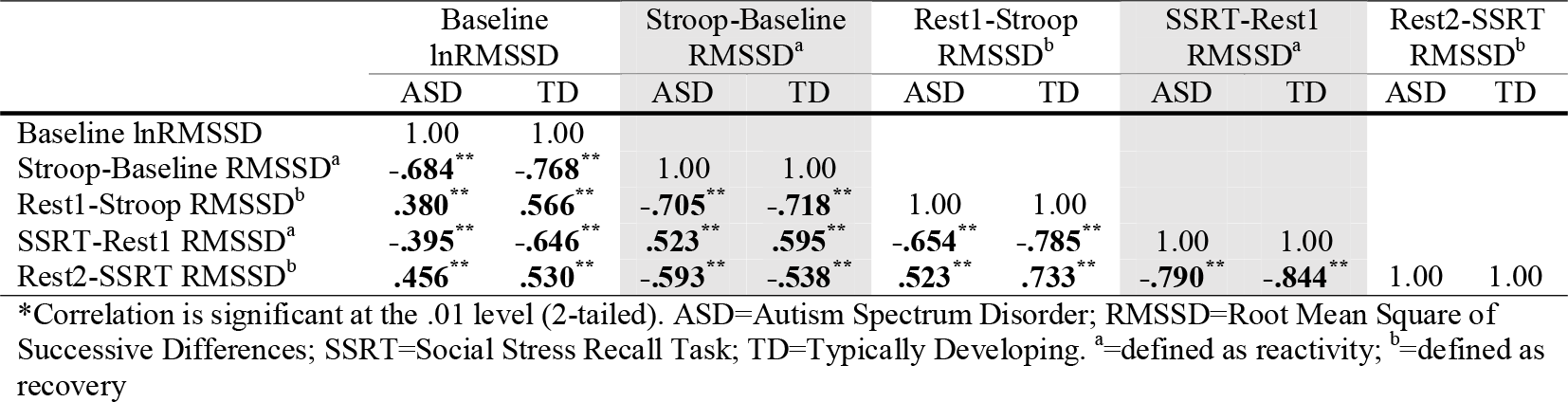
Associations between levels of baseline cardiac vagal modulation and reactivity and recovery levels of cardiac vagal modulation per group as calculated by Spearman’s correlation analysis

The control-analyses using independent samples t-test for baseline HF-HRV revealed lower cardiac vagal modulation in adolescents with ASD with respect to their TD peers (t(92)=2.362, *p*=.020, *d*=.487). Mann-Whitney U-tests also revealed lower HF-HRV values in adolescents with ASD for Rest1 (*U*=830, z=-2.076, *p*=.38, r=.214) and Rest2 (*U*=737, z=-2.779, *p*=.005, r=.287).

### Relationship of cardiac vagal modulation with psychosocial functioning and self-perceived stress

The results are presented per group in Table 4. [Please insert Table 4 here] In adolescents with ASD, reduced baseline levels of cardiac vagal modulation were weakly correlated to higher severity of autism characteristics (i.e., indexed by higher scores on several subscales concerning autism characteristics) and to some parent-reported behavioral subscales (*p*<.05). Reduced reactivity levels, defined as more withdrawal of cardiac vagal modulation during the stress-provoking tasks, were weak to moderately correlated to higher severity of autism characteristics and several parent- and self-reported behavioral subscales. Higher self-reported levels of anxiety and stress in adolescents with ASD were also associated with reduced cardiac vagal reactivity levels (p<.05). Faster stress recovery, defined as an increase of cardiac vagal modulation after the stressor, was weak to moderately correlated to reduced severity of autism characteristics, self- and parent reported behavioral symptoms as well as self-reported levels of stress and anxiety in adolescents with ASD.

**Table 4.** Associations of cardiac vagal modulation and self- and parent reported outcomes on psychosocial functioning per group as calculated by Pearson and Spearman’s correlation analysis

|  | Baseline lnRMSSD |  | Stroop-Baseline RMSSD<sup>a</sup> |  | Rest1-Stroop RMSSD<sup>b</sup> |  | SSRT-Rest1 RMSSD<sup>a</sup> |  | Rest2-SSRT RMSSD<sup>b</sup> |  |
| --- | --- | --- | --- | --- | --- | --- | --- | --- | --- | --- |
|  | ASD | TD | ASD | TD | ASD | TD | ASD | TD | ASD | TD |
| SRS Social Cognition | -.180 | -.157 | **.289\*** | -.047 | **-.400\*\*** | -.028 | **.418\*** | .140 | **-.315\*** | -.026 |
| SRS Social Communication | -.198 | -.176 | **.363\*** | .157 | **-.505\*\*** | -.148 | **.472\*\*** | .073 | **-.347\*** | -.074 |
| SRS Social Motivation | **-.309\*** | -.140 | **.389\*\*** | .036 | **-.407\*\*** | -.001 | **.377\*\*** | .117 | -.264 | -.038 |
| SRS Autistic Preoccupations | -.165 | -.105 | **.297\*** | -.121 | **-.339\*** | .105 | **.300\*** | .077 | -.275 | -.040 |
| SRS Total Score | -.210 | -.229 | **.365\*** | .123 | **-.472\*\*** | -.098 | **.454\*\*** | .105 | **-.343\*** | -.045 |
| RBS Stereotyped Behavior | -.144 | -.177 | .164 | .031 | -.108 | .163 | **.309\*** | .022 | -.219 | .042 |
| RBS Compulsive Behavior | -.131 | -.019 | .251 | -.185 | -.194 | .183 | **.325\*** | -.111 | **-.293\*** | .118 |
| RBS Ritualistic Behavior | **-.396\*\*** | -.023 | **.515\*\*** | -.042 | **-.363\*** | .100 | **.379\*\*** | -.052 | **-.330\*** | -.016 |
| RBS Sameness Behavior | **-.365\*** | .037 | **.421\*\*** | -.080 | **-.343\*** | .125 | **.385\*\*** | -.164 | -.272 | .233 |
| RBS Total Score | **-.376\*\*** | -.033 | **.404\*\*** | -.188 | **-.303\*** | .206 | **.394\*\*** | -.167 | **-.327\*** | .171 |
| SDQ-P Emotional<sup>c</sup> | **-.325\*** | -.171 | **.341\*** | -.024 | **-.324\*** | -.027 | **.435\*\*** | .225 | **-.420\*\*** | -.143 |
| SDQ-P Hyperactivity<sup>c</sup> | .019 | **-.302\*** | .013 | .236 | -.162 | **-.303\*** | .250 | **.340\*** | **-.314\*** | -.193 |
| SDQ-P Peer Problems<sup>c</sup> | -.175 | -.054 | **.325\*** | -.155 | **-.383\*\*** | .045 | .279 | .027 | -.221 | .008 |
| SDQ-P Total Score<sup>c</sup> | -.243 | -.221 | **.324\*** | .095 | **-.448\*\*** | -.096 | **.491\*\*** | .190 | **-.472\*\*** | -.115 |
| SDQ-S Emotional | -.219 | .032 | .204 | .050 | **-.314\*** | .067 | **.409\*\*** | -.015 | -.262 | .181 |
| SDQ-S Hyperactivity | .060 | -.009 | -.096 | .181 | -.020 | -.211 | **.338\*** | .081 | **-.395\*\*** | .064 |
| SDQ-S Prosocial Behavior | -.011 | -.023 | **-.327\*** | .058 | .210 | -.200 | .016 | .258 | .225 | -.124 |
| SDQ-S Total Score | -.127 | -.039 | .156 | .136 | **-.290\*** | -.062 | **.409\*\*** | .052 | **-.300\*** | .129 |
| DASS Anxiety | -.116 | -.242 | .094 | .246 | **-.304\*** | -.050 | **.385\*\*** | .032 | -.237 | .155 |
| DASS Stress | -.090 | -.134 | .169 | .125 | **-.340\*** | .048 | **.351\*** | .091 | -.259 | .102 |
| DASS Total Score | -.097 | -.231 | .114 | .242 | -.271 | -.064 | **.316\*** | .126 | -.158 | .066 |
| PSS | -.157 | -.235 | .134 | .151 | -.285 | .049 | **.314\*** | .069 | -.197 | .086 |
| Sensory hyper responsivity | -.134 | .109 | .240 | -.127 | **-.321\*** | -.141 | .280 | .194 | -.261 | -.095 |

\*Correlation is significant at .05 level (2 tailed); \*\* Correlation is significant at .01 level (2 tailed); <sup>a</sup>=defined as reactivity; <sup>b</sup>=defined as recovery; <sup>c</sup>=1 missing value for adolescent with ASD. ASD=Autism Spectrum Disorder; DASS=Depression, Anxiety and Stress Scale; PSS=Perceived Stress Scale; RBS=Repetitive Behavior Scale; RMSSD=Root Mean Square of Successive Differences; SDQ=Strengths and Difficulties Questionnaire; SRS=Social Responsiveness Scale; TD=Typically Developing

For TD adolescents, higher baseline, reactivity and recovery levels of cardiac vagal modulation were weakly correlated to less parent-reported hyperactivity (resp. *r*_*s*_=-.302, *r*_*s*_=.340, *r*_*s*_=-.303, *p*<.05) as assessed with the SDQ.

### Physiological reactivity and cortisol

#### Heart rate

Repeated measures ANOVA with group as a between-group variable and phase as a within-group variable revealed a significant effect on heart rate over time (F(2.925,269.085)=69.304, *p*<.001, η^2^_p_=.430) but no significant phase x group interaction (*p*>.05). However, a main effect of group was identified, indicating that, irrespective of phase, adolescents with ASD had a significantly higher heart rate with respect to their TD peers (F(1,92)=5.394, *p*=.022, η^2^_p_=.055) (see Figure 2). [Please insert Fig. 2 here]

**Fig. 2.**
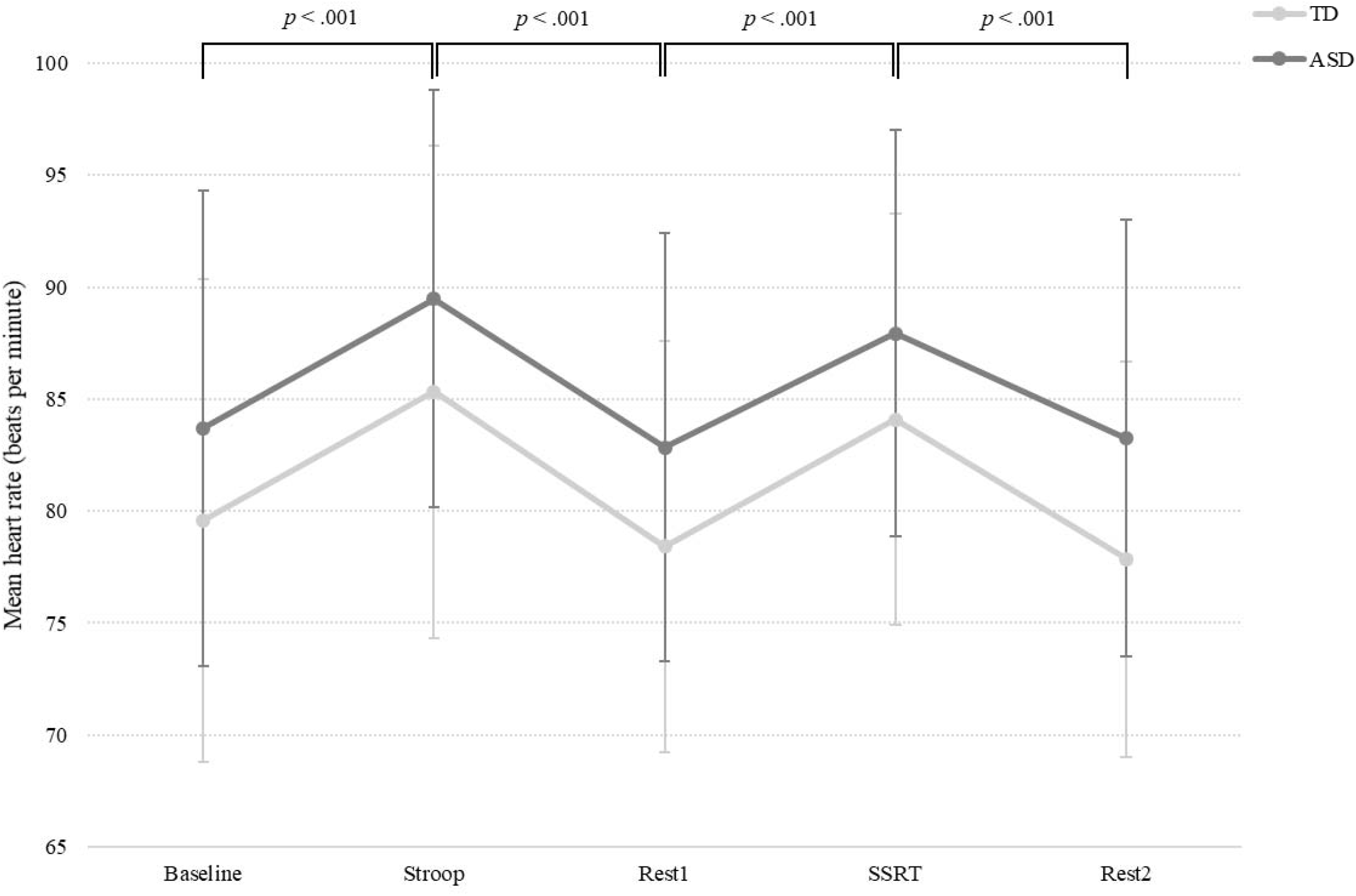
Change in heart rate frequency during the stress-provoking assessment. Note: ASD=Autism Spectrum Disorder, SSRT=Social Stress Recall Task, TD=Typically Developing

#### Breathing frequency

Repeated measures ANOVA with group as a between-group variable and phase as a within-group variable revealed a significant effect on breathing frequency over time (F(3.390,311.848)=70.461, *p*<.001, η^2^_p_ =.434) and a significant phase x group interaction effect (F(3.390,311.848)=4.451, *p*=.003, η^2^_p_=.046). The interaction effect revealed a change of breathing frequency from the first resting period towards the end of the assessment between both groups where adolescents with ASD showed less task-adapted change in breathing frequency. No significant main effect of group was evident. (*p*>.05) (see Figure 3). [Please insert Fig. 3 here]

**Fig. 3.**
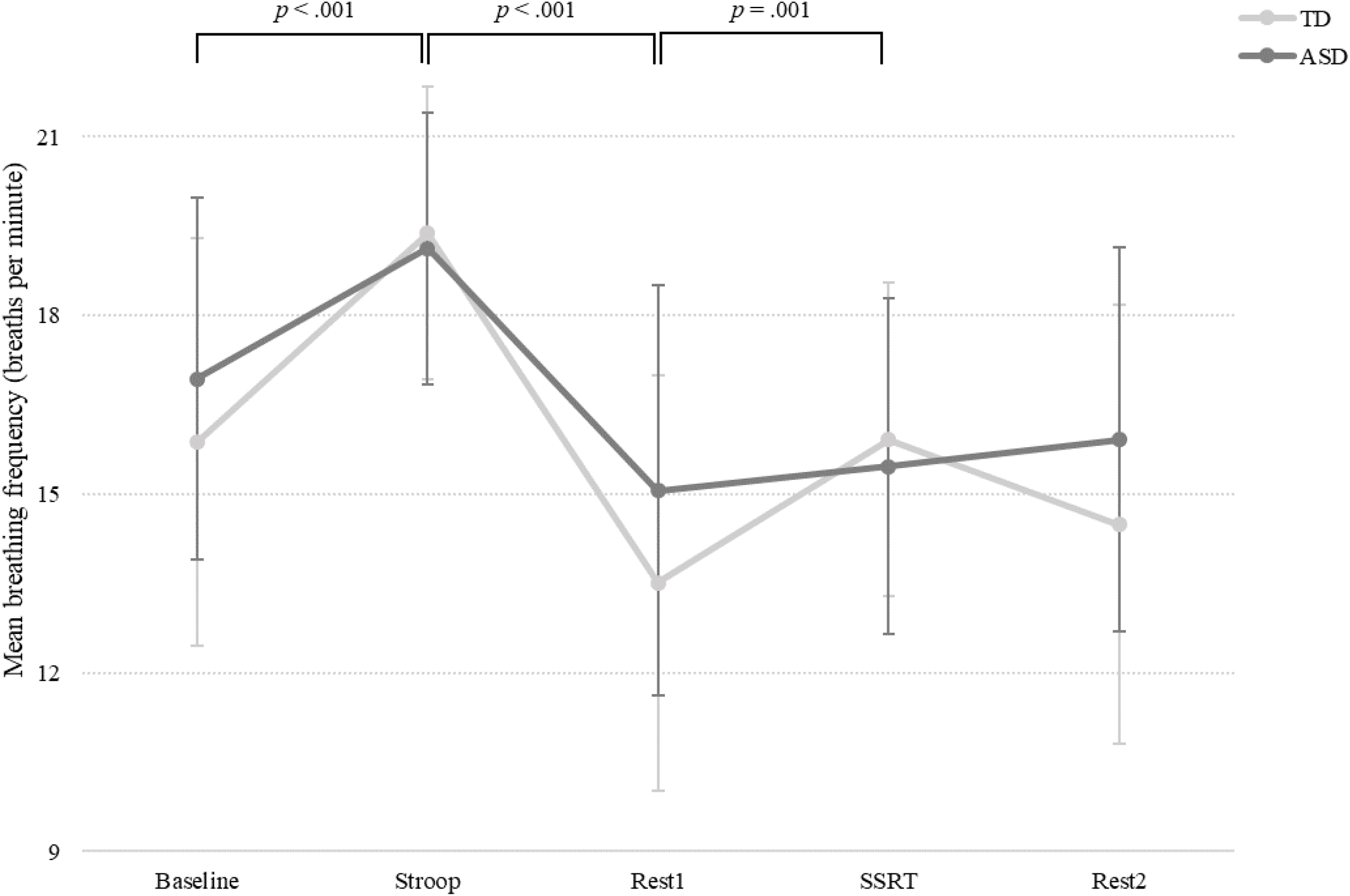
Change in breathing frequency during the stress-provoking assessment. Note: ASD=Autism Spectrum Disorder, SSRT=Social Stress Recall Task, TD=Typically Developing

#### Cortisol

Repeated measures ANOVA with group as a between-group variable and phase as a within-group variable revealed a significant effect of phase (F(1.260,112.144)=15.443, *p*<.001, η^2^_p_=.148) but no significant phase x group interaction effect (*p*>.05). In addition, no significant main effect of group was identified, indicating that the levels of cortisol did not differ between the two groups (*p*>.05; see Figure 4). [Please insert Fig. 4 here] Pearson and Spearman’s correlation analyses revealed no significant correlations between baseline cortisol levels and the level of cardiac vagal modulation at baseline for adolescents with ASD and their TD peers, respectively (*p*>.05).

**Fig. 4.**
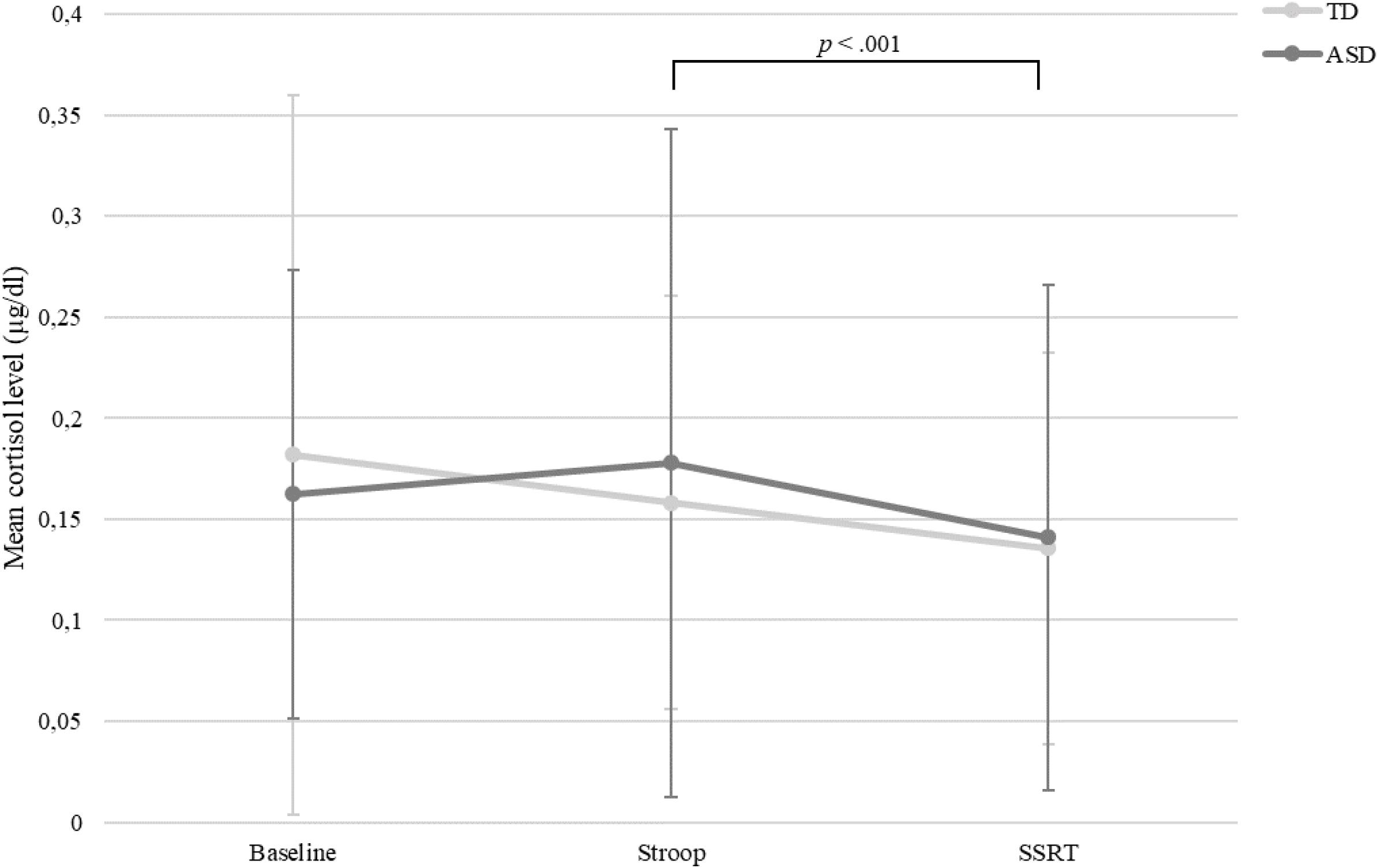
Change in cortisol level during the stress-provoking assessment. Note: ASD=Autism Spectrum Disorder, SSRT=Social Stress Recall Task, TD=Typically Developing

## Discussion

The purpose of this cross-sectional study was to compare the level of cardiac vagal modulation as well as the reactivity and recovery levels of cardiac vagal modulation during baseline and two stressors (social and cognitive) between adolescents with and without ASD. Furthermore, associations with clinical and behavioral indices were explored in addition to changes in cortisol levels during the assessment. The findings of this study support the first hypothesis stating that adolescents with ASD display lower levels of cardiac vagal modulation during baseline as well as during stress-induction with respect to their TD peers. However, the overall extent of stressor-induced changes, in terms of cardiac vagal reactivity and recovery, was similar across groups, thus not supporting the second hypothesis. The third hypothesis was partially confirmed for adolescents with ASD as the study findings indicated some associations between the level of cardiac vagal modulation and self- and parent-reported measures of psychosocial functioning and autism characteristics. In contrast, no significant associations were found between baseline cortisol levels and cardiac vagal modulation in both groups. Furthermore, the cortisol levels of adolescents with and without ASD did not differ during baseline and both stress-provoking tasks, with a significant decrease in the cortisol level of both groups during the SSRT. Finally, both heart rate and breathing frequency changed significantly over time, indicating a consistently higher heart rate in adolescents with ASD.

The lower values of cardiac vagal modulation during baseline and stress in adolescents with ASD found in this study are in line with previously reported findings of lower parasympathetic activity in children and adolescents with ASD (Benevides &Lane, 2015; Cheng et al., 2020; Condy et al., 2019; Makris et al., 2022). Importantly, this study revealed similar reactivity and recovery levels towards the social stressor and cognitive stressor in adolescents with and without ASD, which is in line with the results of Edmiston et al. (2016) and Muscatello et al. (2021), who examined the cardiac vagal reactivity in the same groups using a social stressor. The studies of Levine et al. (2012) and Lory et al. (2020) also demonstrated similar results for a cognitive stressor and a non-social auditory attentional task, respectively. As in this study, they did not find a significant interaction effect between phase of the assessment and group (ASD or TD).

Since two stress-provoking tasks were used, a decreased activity of the parasympathetic nervous system and thus cardiac vagal modulation was expected in response to these tasks. In addition, higher baseline values of cardiac vagal modulation were expected to induce larger levels of reactivity and recovery, based on the ‘law of initial values’ (Wilder, 1967) and as suggested by Patriquin et al. (2011). Consistent with this hypothesis, weak to strong correlations were present implicating that higher baseline levels of cardiac vagal modulation led to larger reactivity and recovery levels in both groups. However, the correlations in the TD group were stronger in contrast to those of the adolescents with ASD. A possible explanation might be that the significantly higher baseline cardiac vagal modulation in TD adolescents, implicates that larger decreases in cardiac vagal modulation were possible during the stress-provoking tasks and, logically, larger increases of this parameter during rest.

Overall, cardiac vagal modulation is suggested to be an important biomarker for social behavior, self-regulatory behavior and emotion-regulation (Beauchaine, 2015; Benevides &Lane, 2015; Condy et al., 2019; Edmiston et al., 2016; Makris et al., 2022; Morrish, 2019; Neuhaus et al., 2016; Patriquin et al., 2014; Patriquin et al., 2015; Patriquin et al., 2011). In line with this notion, the current study showed that higher baseline levels of cardiac vagal modulation in adolescents with ASD were associated with less problems in social motivation, less ritualistic behavior and insistence on sameness behavior in addition to a lower RBS-R total score and less parent-reported emotional problems. Although the associations were weak, these findings build upon the results of Condy et al. (2019), who reviewed the few studies linking baseline cardiac vagal modulation to repetitive and restrictive behaviors. They suggested using the RBS-R to have a more comprehensive assessment of repetitive and restrictive behaviors, as was done in this study. With respect to social functioning, the results of Patriquin et al. (2011) were partially replicated as higher levels of baseline cardiac vagal modulation are indeed associated with better social functioning. Importantly, this study used the SRS-2 as an outcome measure for social functioning whereas the findings of Patriquin et al. (2011) were based on observational data of social behavior. Finally, lower baseline levels of cardiac vagal modulation have been associated with clinical problems characterized by difficulties with self- and emotion regulation (Beauchaine, 2015). Although only parent-reported emotional problems of the adolescents with ASD were significantly correlated with baseline cardiac vagal modulation, this finding builds upon this previous research. Of note, none of the self-reported outcome measures correlated with baseline cardiac vagal modulation. Although criticism is frequently expressed regarding self-reports in individuals with ASD, Arora et al. (2021) plea for the inclusion of these self-reports as emotional experiences of individuals with ASD can sometimes be hard for parents to observe.

As suggested by Porges’ Polyvagal Theory and demonstrated in previous research, larger differences in cardiac vagal modulation in response to environmental demands (such as a stressor) would reflect more appropriate social behavior and self-regulation of behavior and emotions (Condy et al., 2019; Edmiston et al., 2016; Porges, 2011). This study supports previous research, as significant associations were found between psychosocial functioning (using both self- and parent reported measures) and levels of cardiac vagal reactivity and recovery. First, up until now, the link between the level of cardiac vagal reactivity and recovery and the presence of repetitive and restrictive behaviors in adolescents with ASD remained unexplored as prior studies focused on baseline cardiac vagal modulation only (Condy et al., 2019). In this study, larger decreases in cardiac vagal reactivity (change from baseline or rest in response to both stressors) were related to lower scores on the RBS-R and thus less repetitive and restrictive behaviors. The same was true for cardiac vagal recovery values during the resting periods after both stressors although, in this case, larger increases in cardiac vagal modulation were associated with less repetitive and restrictive behaviors. Second, more cardiac vagal reactivity and recovery was also associated with less social problems (as assessed with the SRS-2) and emotional problems (as assessed with the SDQ). Important to note is that not all subscales yielded significant results but all total scores from the questionnaires did. Third, lower levels of cardiac vagal reactivity and recovery were associated with higher levels of self-reported anxiety and stress (assessed with the DASS and PSS), supporting the relation of cardiac vagal modulation with emotion-regulation. Fourth, an inverse relationship between higher cardiac vagal modulation and less sensory hyperresponsivity was demonstrated for cardiac vagal reactivity during the Stroop task, but not during the SSRT, which partially supports the findings of Matsushima et al. (2016).

Important to note are the group-based differences between measures of psychosocial functioning and baseline cardiac vagal modulation as well as with cardiac vagal reactivity and recovery levels. Only one significant association was found between lower levels of parent-reported hyperactivity in TD adolescents and higher levels of baseline cardiac vagal modulation and reactivity and recovery levels. However, this study used the SDQ, which results in a categorical outcome, which may have led towards less differentiation for the TD adolescents and thus may have hampered appropriate correlation analyses with cardiac vagal modulation.

Since both the ANS and the HPA-axis are involved in the stress response, the association between cardiac vagal modulation and cortisol level at baseline was examined. However, no significant correlations were found in contrast to Corbett et al. (2019), who did find that higher levels of cardiac vagal modulation were associated with lower levels of cortisol in their sample of children with ASD but not in their TD group. They explained this finding by stating that the variability of cardiac vagal modulation in their TD group was lower than that of the children with ASD. In this study, a similar variability was found for cardiac vagal modulation in both groups whereas the direction of the differences between the groups in variability of cortisol levels was different for baseline, Stroop and SSRT, respectively. In addition, the baseline cortisol levels were collected before the actual baseline measurement during which cardiac vagal modulation was measured. Thus, a more appropriate timing might have been to perform the baseline cortisol measurement at 20 minutes after the actual baseline measurement of the stress-provoking assessment, as was performed for the Stroop and SSRT. Furthermore, no significant group-based differences in cortisol were found in this study across the assessment in contrast with the significantly lower levels of cardiac vagal modulation in adolescents with ASD. This is counterintuitive given that these lower cardiac vagal modulation levels may imply a heightened state of arousal and would thus result in higher cortisol values. As depicted in Figure 4, cortisol levels in adolescents with ASD tended to increase during the Stroop task, although it did not reach the level of significance. The Stroop task has been used previously in children with ASD to investigate the level of autonomic response but not the response of the HPA axis (Kushki et al., 2013). Up until now, no studies have used the Stroop task as a cognitive stressor to induce an HPA axis response, making it difficult to compare these results with respect to other studies. However, other cognitive tasks have been used in child populations with various psychiatric disorders, including ASD, such as an academic performance test (Anesiadou et al., 2021) and a continuous performance task (Jansen et al., 1999) but these also did not trigger an HPA axis response. The SSRT on the other hand, was included as a social stressor. In the study of Bishop-Fitzpatrick et al. (2017), adults with ASD demonstrated an increase of cortisol levels in response to the SSRT. However, this study could not replicate these findings in adolescents with and without ASD as cortisol levels decreased in response to the SSRT. Furthermore, TD adolescents showed lower cortisol levels during both the Stroop task and the SSRT. In sum, the mild nature of the stressors and the absence of a physiologic threat may have contributed to the absence of a significant increase in cortisol for both groups.

The consistent higher heart rate in adolescents with ASD is a result of the lower influence of cardiac vagal modulation in this study sample. The vagal nerve is hypothesized to work as a ‘vagal brake’ on heart rate by decreasing the high intrinsic rate of the heart’s pacemaker, when activated (Porges, 2011). Building upon this reasoning, lower influence of the vagal nerve (or lower cardiac vagal modulation) would result in a higher heart rate, as was found in this study. The significant differences in breathing frequency across the assessment can be logically explained by the nature of the tasks since baseline and resting periods were characterized by sitting calmly without talking while both stressors were verbal tasks. The significant interaction effect on the breathing frequency may be explained by the hypothesis that adolescents with ASD were less able to stabilize their breathing frequency after a stressor.

Overall, lower cardiac vagal modulation is not uniquely found in individuals with ASD but across a variety of psychiatric disorders such as depressive disorders, anxiety disorders, substance use disorders, schizophrenia and personality disorders (Heiss et al., 2021). These disorders frequently co-occur in individuals with ASD (Al-Beltagi, 2021) and thus may have influenced the study findings. Co-occurring disorders were reported for 46% of the adolescents with ASD but the small groups for each of the co-occurring disorders hampered the performance of a reliable covariate analysis. In addition, use of psychotropic medications such as stimulants, antipsychotics and antidepressants have been associated with reductions in cardiac vagal modulation (Thapa et al., 2019). Therefore, the use of psychotropic medication was used as in additional covariate analyses in this study but revealed no significant influence on cardiac vagal modulation (*p*>.05).

### Clinical relevance and future research

As suggested by multiple researchers, interventions based on an upregulation of the activity of the parasympathetic nervous system might induce positive effects on behavior as well (Condy et al., 2019; Heiss et al., 2021; Patriquin et al., 2019). In addition, a better understanding of the functioning of both branches of the ANS, as this study only discussed the parasympathetic branch, and the coordination with the HPA axis might shed a light on the large variability in findings of previous research together with the current study findings. This may ultimately lead towards unravelling possible subtypes of physiological reactivity that are associated with different expressions and severity of autism characteristics (Arora et al., 2021; Beauchaine et al., 2019; Benevides &Lane, 2015; Makris et al., 2022; Morrish, 2019; Muscatello et al., 2022; Taylor &Corbett, 2014). Building upon the Neurovisceral Integration Theory, it would be of great relevance to combine physiological measures with neurological imaging to detect activation or inhibition of specific brain regions during challenging tasks and resting conditions. This way, a complete overview of top-down and bottom-up processes can be mapped with respect to ANS and HPA axis functioning (Arora et al., 2021; Cheng et al., 2020; Condy et al., 2019; Kushki et al., 2014; Patriquin et al., 2019).

## Strengths and limitations

This study includes a rigorously characterized sample of adolescents with and without ASD, a highly standardized assessment, measures of both cardiac vagal modulation and HPA axis functioning and a robust statistical approach. Despite these strengths, there are important limitations that need to be acknowledged.

First, the study sample did not include adolescents with a diagnosis of intellectual disability despite findings suggesting a protective role of IQ leading to higher cardiac vagal modulation in children and adolescents with ASD without intellectual disability (Porges et al., 2013). However, the study design was deemed unsuitable for individuals with a diagnosis of intellectual disability. Therefore, the findings of this study cannot be generalized towards all adolescents with ASD.

Second, the non-significant difference in cardiac vagal modulation between baseline and the Stroop task in both groups might have been caused by the long duration (10 minutes) of the baseline measurement. This timing was based on the recommendations of Quintana et al. (2016), who suggested the inclusion of an acclimatization phase before data collection starts in addition to withholding the instruction that the actual recording has started. However, other researchers suggested the use of a so-called vanilla task, which consists of baseline measurements during which participants listen to music or watch a movie (Jennings et al., 1992). Nevertheless, reluctance has been expressed towards the use of these tasks since reactivity of cardiac vagal modulation may be induced by attention allocations mechanisms (Laborde et al., 2017).

Third, non-significant findings between the measures of depressive symptoms and cardiac vagal modulation are surprising given cumulating evidence of a negative association in other studies (Hamilton &Alloy, 2016). Together with other non-significant findings between cardiac vagal modulation and measures of psychosocial functioning and self-perceived stress in this study, it should be stated that more differentiating measures should be used in the future. As suggested by Beauchaine et al. (2019), nonlinear associations between physiological functioning and psychopathology may be more informative. They state that certain levels of withdrawal of cardiac vagal modulation may be related to adaptive functioning, whereas extreme reactivity levels may be associated with pathology (Beauchaine et al., 2019; Beauchaine et al., 2007). Furthermore, the non-significant correlation between self-reported stress and physiological measures at baseline in individuals with ASD has been discussed in the past by stating that both measures may account for different constructs of stress, which may result in the absence of significant correlations (Thoen et al., 2021). In addition, using a standardized assessment tool for sensory hyper responsivity across multiple sensory modes (Short Sensory Profile; McIntosh et al., 1999) as in previous research (Matsushima et al., 2016) may have been more appropriate.

Fourth, the primary outcome measure in this cross-sectional study is cardiac vagal modulation, which is influenced by numerous factors such as the level of physical activity (Stanley et al., 2013). Therefore, a short questionnaire reporting the number of days and minutes that adolescents spent on moderate to vigorous physical activity was included in this study (Physical Activity Vital Sign; Greenwood et al., 2010). Unfortunately, 24% of the responses were discarded due to unsuspected results leading to the assumption that some adolescents with and without ASD misinterpreted the questions. Researchers using this questionnaire in the future might prefer to fill it out together with the adolescents instead of using an online mode as was done in this study.

## Conclusion

Adolescence is characterized by changes on several developmental domains and is therefore described as a critical transition period with heightened vulnerability towards stressors and the effects of stress (Chrousos, 2009). Findings of this study suggest parasympathetic hypo-activity in adolescents with ASD, although the level of reactivity and recovery was the same as TD peers. This hypo-activity is related to several aspects of psychosocial functioning. In addition, the study findings provided more evidence to consider baseline cardiac vagal modulation as well as its reactivity and recovery levels as a biomarker for social and self-regulatory behavior in adolescents with ASD.

## Data Availability

All data produced in the present study are available upon reasonable request to the authors

## Notes

### Competing Interest Statement

The authors have declared no competing interest.

### Clinical Trial

NCT04628715

### Clinical Protocols

https://doi.org/10.1186/s13063-021-05709-4

### Funding Statement

This work is supported by the Marguerite-Marie Delacroix foundation with grant number RVC/B-472

### Author Declarations

Ethics Committee UPC KU Leuven gave ethical approval for this work (ref. EC2020-541) Ethics Committee Research UZ/KU Leuven gave ethical approval for this work (ref. S64219).

## References

Agorastos, A., Pervanidou, P., Chrousos, G. P., & Kolaitis, G. (2018). Early life stress and trauma: developmental neuroendocrine aspects of prolonged stress system dysregulation. Hormones, 17(4), 507–520. https://doi.org/10.1007/s42000-018-0065-x

Al-Beltagi, M. (2021). Autism medical comorbidities. World journal of clinical pediatrics, 10(3), 15–28. https://doi.org/10.5409/wjcp.v10.i3.15

American Psychiatric Association. (2000). Diagnostic and statistical manual of mental disorders (4, text rev ed.)

American Psychiatric Association. (2013). Diagnostic and statistical manual of mental disorders (5th ed.). American Psychiatric Association. https://doi.org/10.1176/appi.books.9780890425596

Anesiadou, S., Makris, G., Michou, M., Bali, P., Papassotiriou, I., Apostolakou, F., Korkoliakou, P., Papageorgiou, C., Chrousos, G., & Pervanidou, P. (2021). Salivary cortisol and alpha-amylase daily profiles and stress responses to an academic performance test and a moral cognition task in children with neurodevelopmental disorders. Stress and Health, 37(1), 45–59. https://doi.org/10.1002/smi.2971

Arora, I., Bellato, A., Ropar, D., Hollis, C., & Groom, M. J. (2021). Is autonomic function during resting-state atypical in Autism: A systematic review of evidence. Neuroscience & Biobehavioral Reviews, 125, 417–441. https://doi.org/10.1016/j.neubiorev.2021.02.041

Baron-Cohen, S., Hoekstra, R. A., Knickmeyer, R., & Wheelwright, S. (2006). The autism-spectrum quotient (AQ)—adolescent version. Journal of Autism and Developmental Disorders, 36(3), 343. https://doi.org/10.1007/s10803-006-0073-6

Beauchaine, T. P. (2015). Respiratory Sinus Arrhythmia: A Transdiagnostic Biomarker of Emotion Dysregulation and Psychopathology. Current Opinion in Psychology, 3, 43–47. https://doi.org/10.1016/j.copsyc.2015.01.017

Beauchaine, T. P., Bell, Z., Knapton, E., McDonough[Caplan, H., Shader, T., & Zisner, A. (2019). Respiratory sinus arrhythmia reactivity across empirically based structural dimensions of psychopathology: A meta[analysis. Psychophysiology, 56(5), e13329. https://doi.org/10.1111/psyp.13329

Beauchaine, T. P., Gatzke-Kopp, L., & Mead, H. K. (2007). Polyvagal Theory and developmental psychopathology: Emotion dysregulation and conduct problems from preschool to adolescence. Biological Psychology, 74(2), 174–184. https://doi.org/10.1016/j.biopsycho.2005.08.008

Benevides, T. W., & Lane, S. J. (2015). A review of cardiac autonomic measures: considerations for examination of physiological response in children with autism spectrum disorder. Journal of Autism and Developmental Disorders, 45(2), 560–575. https://doi.org/10.1007/s10803-013-1971-z

Bishop-Fitzpatrick, L., Minshew, N. J., Mazefsky, C. A., & Eack, S. M. (2017). Perception of Life as Stressful, Not Biological Response to Stress, is Associated with Greater Social Disability in Adults with Autism Spectrum Disorder. Journal of Autism and Developmental Disorders, 47(1), 1–16. https://doi.org/10.1007/s10803-016-2910-6

Bujnakova, I., Ondrejka, I., Mestanik, M., Visnovcova, Z., Mestanikova, A., Hrtanek, I., Fleskova, D., Calkovska, A., & Tonhajzerova, I. (2016). Autism spectrum disorder is associated with autonomic underarousal. Physiological research, 65. https://doi.org/10.33549/physiolres.933528

Cheng, Y.-C., Huang, Y.-C., & Huang, W.-L. (2020). Heart rate variability in individuals with autism spectrum disorders: A meta-analysis. Neuroscience & Biobehavioral Reviews, 118, 463–471. https://doi.org/10.1016/j.neubiorev.2020.08.007

Chrousos, G. P. (2009). Stress and disorders of the stress system. Nature Reviews Endocrinology, 5(7), 374–381. https://doi.org/10.1038/nrendo.2009.106

Condy, E. E., Scarpa, A., & Friedman, B. H. (2019). Restricted repetitive behaviors in autism spectrum disorder: A systematic review from the neurovisceral integration perspective. Biological Psychology, 148, 107739. https://doi.org/10.1016/j.biopsycho.2019.107739

Constantino, J., & Gruber, C. (2015). Screeningslijst voor autismespectrumstoornissen (SRS-2): Nederlandse bewerking (H. Roeyers, M. Thys, C. Druart, M. D. Schryver, & M. Schittekatte Trans.). Hogrefe Uitgevers B.V. (Original work published in 2012)

Constantino, J. N., & Gruber, C. P. (2012). Social Responsiveness Scale Second Edition (SRS-2): Manual. Western Psychological Services (WPS).

Corbett, B. A., Muscatello, R. A., & Baldinger, C. (2019). Comparing stress and arousal systems in response to different social contexts in children with ASD. Biological Psychology, 140, 119–130. https://doi.org/10.1016/j.biopsycho.2018.12.010

Corbett, B. A., Muscatello, R. A., Kim, A., Patel, K., & Vandekar, S. (2021). Developmental effects in physiological stress in early adolescents with and without autism spectrum disorder. Psychoneuroendocrinology, 125, 105115. https://doi.org/10.1016/j.psyneuen.2020.105115

de Beurs, E., Van Dyck, R., Marquenie, L. A., Lange, A., & Blonk, R. W. B. (2001). De DASS: Een vragenlijst voor het meten van depressie, angst en stress. [The DASS: A questionnaire for the measurement of depression, anxiety, and stress.]. Gedragstherapie, 34(1), 35–53.

Edmiston, E. K., Blain, S. D., & Corbett, B. A. (2017). Salivary cortisol and behavioral response to social evaluative threat in adolescents with autism spectrum disorder. Autism Research, 10(2), 346–358. https://doi.org/10.1002/aur.1660

Edmiston, E. K., Jones, R. M., & Corbett, B. A. (2016). Physiological response to social evaluative threat in adolescents with autism spectrum disorder. Journal of Autism and Developmental Disorders, 46(9), 2992–3005. https://doi.org/10.1007/s10803-016-2842-1

Fiskum, C. (2019). Psychotherapy Beyond All the Words: Dyadic Expansion, Vagal Regulation, and Biofeedback in Psychotherapy. Journal of psychotherapy integration, 29(4), 412–425. https://doi.org/10.1037/int0000174

Foley, P., & Kirschbaum, C. (2010). Human hypothalamus–pituitary–adrenal axis responses to acute psychosocial stress in laboratory settings. Neuroscience & Biobehavioral Reviews, 35(1), 91–96. https://doi.org/10.1016/j.neubiorev.2010.01.010

Friedman, B. H. (2007). An autonomic flexibility–neurovisceral integration model of anxiety and cardiac vagal tone. Biological Psychology, 74(2), 185–199. https://doi.org/10.1016/j.biopsycho.2005.08.009

Goodman, R. (2001). Psychometric properties of the strengths and difficulties questionnaire. Journal of the American Academy of Child & Adolescent Psychiatry, 40(11), 1337–1345. https://doi.org/10.1097/00004583-200111000-00015

Greenwood, J., Joy, E., & Stanford, J. (2010). The Physical Activity Vital Sign: a primary care tool to guide counseling for obesity. Journal of Physical Activity and Health, 7, 571–576. https://doi.org/10.1123/jpah.7.5.571

Grossman, P., Stemmler, G., & Meinhardt, E. (1990). Paced Respiratory Sinus Arrhythmia as an Index of Cardiac Parasympathetic Tone During Varying Behavioral Tasks. Psychophysiology, 27(4), 404–416. https://doi.org/10.1111/j.1469-8986.1990.tb02335.x

Hamilton, J. L., & Alloy, L. B. (2016). Atypical reactivity of heart rate variability to stress and depression across development: Systematic review of the literature and directions for future research. Clinical Psychology Review, 50, 67–79. https://doi.org/10.1016/j.cpr.2016.09.003

Heiss, S., Vaschillo, B., Vaschillo, E. G., Timko, C. A., & Hormes, J. M. (2021). Heart rate variability as a biobehavioral marker of diverse psychopathologies: A review and argument for an “ideal range”. Neuroscience & Biobehavioral Reviews, 121, 144–155. https://doi.org/10.1016/j.neubiorev.2020.12.004

Hollocks, M. J., Howlin, P., Papadopoulos, A. S., Khondoker, M., & Simonoff, E. (2014). Differences in HPA-axis and heart rate responsiveness to psychosocial stress in children with autism spectrum disorders with and without co-morbid anxiety. Psychoneuroendocrinology, 46, 32–45. https://doi.org/10.1016/j.psyneuen.2014.04.004

Jansen, L. M. C., Gispen-de Wied, C. C., Jansen, M. A., van der Gaag, R.-J., Matthys, W., & van Engeland, H. (1999). Pituitary–adrenal reactivity in a child psychiatric population: salivary cortisol response to stressors. European Neuropsychopharmacology, 9(1), 67–75. https://doi.org/10.1016/S0924-977X(98)00003-0

Jennings, J. R., Kamarck, T., Stewart, C., Eddy, M., & Johnson, P. (1992). Alternate cardiovascular baseline assessment techniques: vanilla or resting baseline. Psychophysiology, 29(6), 742–750. https://doi.org/10.1111/j.1469-8986.1992.tb02052.x

Kushki, A., Brian, J., Dupuis, A., & Anagnostou, E. (2014). Functional autonomic nervous system profile in children with autism spectrum disorder. Molecular Autism, 5(1), 39. https://doi.org/10.1186/2040-2392-5-39

Kushki, A., Drumm, E., Mobarak, M. P., Tanel, N., Dupuis, A., Chau, T., & Anagnostou, E. (2013). Investigating the autonomic nervous system response to anxiety in children with autism spectrum disorders. PLoS ONE, 8(4). https://doi.org/10.1371/journal.pone.0059730

Laborde, S., Mosley, E., & Thayer, J. F. (2017). Heart Rate Variability and Cardiac Vagal Tone in Psychophysiological Research - Recommendations for Experiment Planning, Data Analysis, and Data Reporting. Frontiers in Psychology, 8, 213. https://doi.org/10.3389/fpsyg.2017.00213

Lam, K. S., & Aman, M. G. (2007). The Repetitive Behavior Scale-Revised: independent validation in individuals with autism spectrum disorders. Journal of Autism and Developmental Disorders, 37(5), 855–866. https://doi.org/10.1007/s10803-006-0213-z

Lehrer, P., Kaur, K., Sharma, A., Shah, K., Huseby, R., Bhavsar, J., & Zhang, Y. (2020). Heart Rate Variability Biofeedback Improves Emotional and Physical Health and Performance: A Systematic Review and Meta Analysis. Applied Psychophysiology and Biofeedback, 45(3), 109–129. https://doi.org/10.1007/s10484-020-09466-z

Levine, T. P., Sheinkopf, S. J., Pescosolido, M., Rodino, A., Elia, G., & Lester, B. (2012). Physiologic arousal to social stress in children with autism spectrum disorders: a pilot study. Research in Autism Spectrum Disorders, 6(1), 177–183. https://doi.org/10.1016/j.rasd.2011.04.003

Lory, C., Kadlaskar, G., McNally Keehn, R., Francis, A. L., & Keehn, B. (2020). Brief Report: Reduced Heart Rate Variability in Children with Autism Spectrum Disorder. Journal of Autism and Developmental Disorders, 50(11), 4183–4190. https://doi.org/10.1007/s10803-020-04458-8

Lydon, S., Healy, O., Reed, P., Mulhern, T., Hughes, B. M., & Goodwin, M. S. (2016). A systematic review of physiological reactivity to stimuli in autism. Developmental Neurorehabilitation, 19(6), 335–355. https://doi.org/10.3109/17518423.2014.971975

Makris, G., Agorastos, A., Chrousos, G. P., & Pervanidou, P. (2022). Stress System Activation in Children and Adolescents With Autism Spectrum Disorder. Frontiers in Neuroscience, 15. https://doi.org/10.3389/fnins.2021.756628

Malik, M. (1996). Heart rate variability: Standards of measurement, physiological interpretation, and clinical use: Task force of the European Society of Cardiology and the North American Society for Pacing and Electrophysiology. Annals of Noninvasive Electrocardiology, 1(2), 151–181.

Matsushima, K., Matsubayashi, J., Toichi, M., Funabiki, Y., Kato, T., Awaya, T., & Kato, T. (2016). Unusual sensory features are related to resting-state cardiac vagus nerve activity in autism spectrum disorders. Research in Autism Spectrum Disorders, 25, 37–46. https://doi.org/10.1016/j.rasd.2015.12.006

McIntosh, D., Miller, L., Shyu, V., & Dunn, W. (1999). Overview of the short sensory profile (SSP). The sensory profile: Examiner’s manual, 59–73.

Morrish, L., Chin, T.C., Rickard, N., Sigley-Taylor, P., & Vella-Brodrick, D.. (2019). The role of physiological and subjective measures of emotion regulation in predicting adolescent wellbeing.International Journal of Wellbeing. International Journal of Wellbeing, 9(2), 66–89. https://doi.org/10.5502/ijw.v9i2.730

Muscatello, R. A., Pachol, A., Romines, A., Smith, I., & Corbett, B. A. (2022). Development and Parasympathetic Regulation in Male and Female Adolescents with Autism Spectrum Disorder: A Two-Timepoint Longitudinal Study. Journal of Autism and Developmental Disorders. https://doi.org/10.1007/s10803-022-05664-2

Muscatello, R. A., Vandekar, S. N., & Corbett, B. A. (2021). Evidence for decreased parasympathetic response to a novel peer interaction in older children with autism spectrum disorder: a case-control study. Journal of Neurodevelopmental Disorders, 13(1), 6. https://doi.org/10.1186/s11689-020-09354-x

Neuhaus, E., Bernier, R. A., & Beauchaine, T. P. (2016). Children with autism show altered autonomic adaptation to novel and familiar social partners. Autism Research, 9(5), 579–591. https://doi.org/10.1002/aur.1543

Patriquin, M. A., Hartwig, E. M., Friedman, B. H., Porges, S. W., & Scarpa, A. (2019). Autonomic response in autism spectrum disorder: Relationship to social and cognitive functioning. Biological Psychology, 145, 185–197. https://doi.org/10.1016/j.biopsycho.2019.05.004

Patriquin, M. A., Lorenzi, J., Scarpa, A., & Bell, M. A. (2014). Developmental trajectories of respiratory sinus arrhythmia: Associations with social responsiveness. Developmental Psychobiology, 56(3), 317–326. http://doi.org/10.1002/dev.21100

Patriquin, M. A., Lorenzi, J., Scarpa, A., Calkins, S. D., & Bell, M. A. (2015). Broad implications for respiratory sinus arrhythmia development: Associations with childhood symptoms of psychopathology in a community sample. Developmental Psychobiology, 57(1), 120–130. https://doi.org/10.1002/dev.21269

Patriquin, M. A., Scarpa, A., Friedman, B. H., & Porges, S. W. (2011). Respiratory sinus arrhythmia: a marker for positive social functioning and receptive language skills in children with autism spectrum disorders. Developmental Psychobiology, 55(2), 101–112. https://doi.org/10.1002/dev.21002

Porges, S. W. (1995). Cardiac vagal tone: a physiological index of stress. Neurosci Biobehav Rev, 19(2), 225–233. https://doi.org/10.1016/0149-7634(94)00066-a

Porges, S. W. (2011). The polyvagal theory: Neurophysiological foundations of emotions, attachment, communication, and self-regulation. W W Norton & Co.

Porges, S. W., Macellaio, M., Stanfill, S. D., McCue, K., Lewis, G. F., Harden, E. R., Handelman, M., Denver, J., Bazhenova, O. V., & Heilman, K. J. (2013). Respiratory sinus arrhythmia and auditory processing in autism: Modifiable deficits of an integrated social engagement system? International Journal of Psychophysiology, 88(3), 261–270. https://doi.org/10.1016/j.ijpsycho.2012.11.009

Quintana, D., Alvares, G. A., & Heathers, J. (2016). Guidelines for Reporting Articles on Psychiatry and Heart rate variability (GRAPH): recommendations to advance research communication. Translational psychiatry, 6(5), e803. https://doi.org/10.1038/tp.2016.73

Richman, L., Bennett, G., Pek, J., Siegler, I., & Williams, R. (2007). Discrimination, dispositions, and cardiovascular responses to stress. Health Psychology, 26, 675–683. https://doi.org/10.1037/0278-6133.26.6.675

Sapolsky, R. M., Romero, L. M., & Munck, A. U. (2000). How Do Glucocorticoids Influence Stress Responses? Integrating Permissive, Suppressive, Stimulatory, and Preparative Actions*. Endocrine Reviews, 21(1), 55–89. https://doi.org/10.1210/edrv.21.1.0389

Stanley, J., Peake, J. M., & Buchheit, M. (2013). Cardiac Parasympathetic Reactivation Following Exercise: Implications for Training Prescription. Sports Medicine, 43(12), 1259–1277. https://doi.org/10.1007/s40279-013-0083-4

Stroop, J. R. (1935). Studies of interference in serial verbal reactions. Journal of experimental psychology, 18(6), 643. https://doi.org/10.1037/h0054651

Taylor, J. L., & Corbett, B. A. (2014). A review of rhythm and responsiveness of cortisol in individuals with autism spectrum disorders. Psychoneuroendocrinology, 49, 207–228. https://doi.org/10.1016/j.psyneuen.2014.07.015

Thapa, R., Alvares, G. A., Zaidi, T. A., Thomas, E. E., Hickie, I. B., Park, S. H., & Guastella, A. J. (2019). Reduced heart rate variability in adults with autism spectrum disorder. Autism Research, 12(6), 922–930. http://doi.org/10.1002/aur.2104

Thayer, J. F., & Lane, R. D. (2000). A model of neurovisceral integration in emotion regulation and dysregulation. Journal of Affective Disorders, 61(3), 201–216. https://doi.org/10.1016/S0165-0327(00)00338-4

Thayer, J. F., & Lane, R. D. (2009). Claude Bernard and the heart–brain connection: Further elaboration of a model of neurovisceral integration. Neuroscience & Biobehavioral Reviews, 33(2), 81–88. https://doi.org/10.1016/j.neubiorev.2008.08.004

Thoen, A., Steyaert, J., Alaerts, K., & Van Damme, T. (2021). Evaluating the potential of respiratory-sinus-arrhythmia biofeedback for reducing physiological stress in adolescents with autism: study protocol for a randomized controlled trial. Trials, 22(1), 730. https://doi.org/10.1186/s13063-021-05709-4

Thoen, A., Steyaert, J., Alaerts, K., Evers, K., Van Damme, T. (2021). A Systematic Review of Self-Reported Stress Questionnaires in People on the Autism Spectrum. Review Journal of Autism and Developmental Disorders. https://doi.org/10.1007/s40489-021-00293-4

Thomas, B. L., Claassen, N., Becker, P., & Viljoen, M. (2019). Validity of Commonly Used Heart Rate Variability Markers of Autonomic Nervous System Function. Neuropsychobiology, 78(1), 14–26. https://doi.org/10.1159/000495519

Tonhajzerova, I., Ondrejka, I., Ferencova, N., Bujnakova, I., Grendar, M., Olexova, L. B., Hrtanek, I., & Visnovcova, Z. (2021). Alternations in the Cardiovascular Autonomic Regulation and Growth Factors in Autism. Physiological Research, 70(4), 551–561. https://doi.org/10.33549/physiolres.934662

Van der Ploeg, J. (2013). Stress bij kinderen. Bohn Stafleu van Loghum, onderdeel van Springer Media BV.

Van Hecke, A. V., Lamb, D., Lebow, J., Bal, E., Harden, E., Kramer, A., Denver, J., Bazhenova, O., & Porges, S. W. (2009). Electroencephalogram and Heart Rate Regulation to Familiar and Unfamiliar People in Children with Autism Spectrum Disorders. Child Development, 80(4), 1118–1133. http://doi.org/10.1111/j.1467-8624.2009.01320.x

Wilder, J. F. (1967). Stimulus and response: The law of initial value. Williams & Wilkins. https://doi.org/10.1016/C2013-0-06704-7

