## Supplementary material for "Differences in cardiac vagal modulation and cortisol response in adolescents with and without Autism Spectrum Disorder"

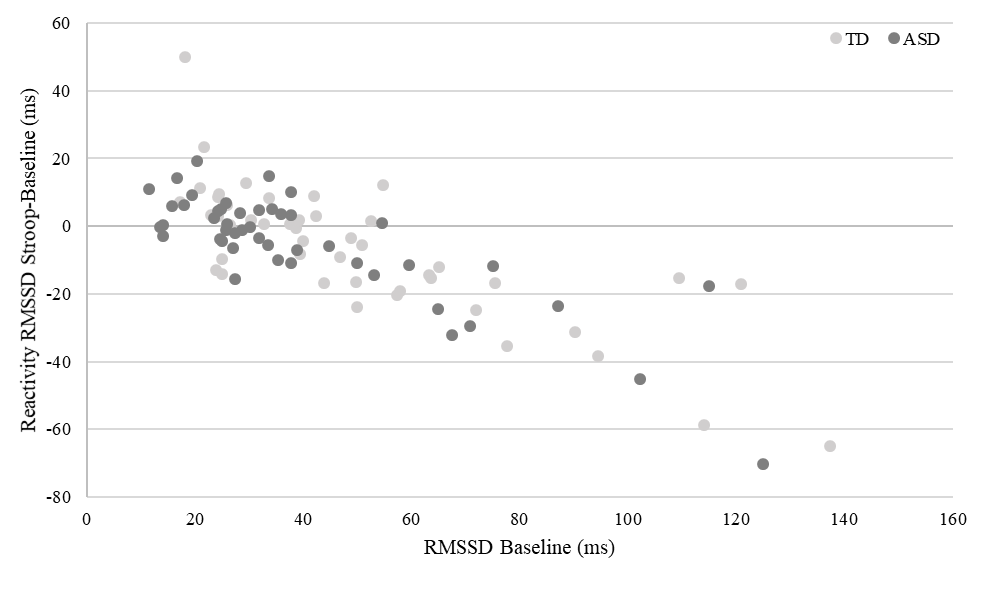

**Supplementary Material A** Association between baseline cardiac vagal modulation and cardiac vagal reactivity during the Stroop task based on Spearman’s correlation analysis (ASD: *r_s_*=-.684; TD: *r_s_*=-.768, *p*<.01). Note: to ease the interpretability of this graph, raw baseline RMSSD values were used

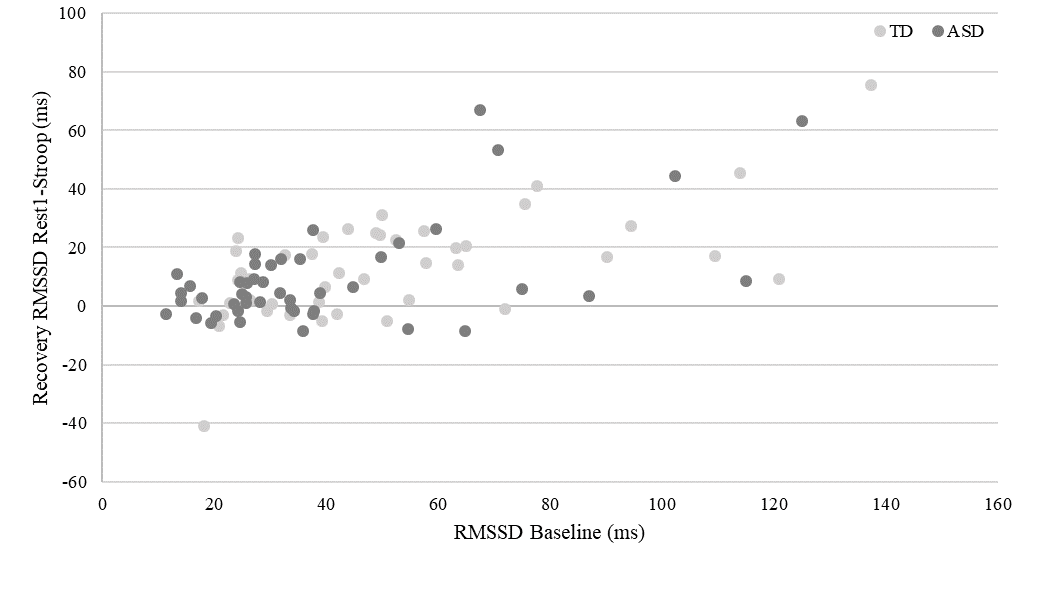

**Supplementary Material B** Association between baseline cardiac vagal modulation and cardiac vagal recovery following the Stroop task based on Spearman’s correlation analysis (ASD: *r_s_*=.380; TD: *r_s_=*.566; *p*<.01). Note: to ease the interpretability of this graph, raw baseline RMSSD values were used

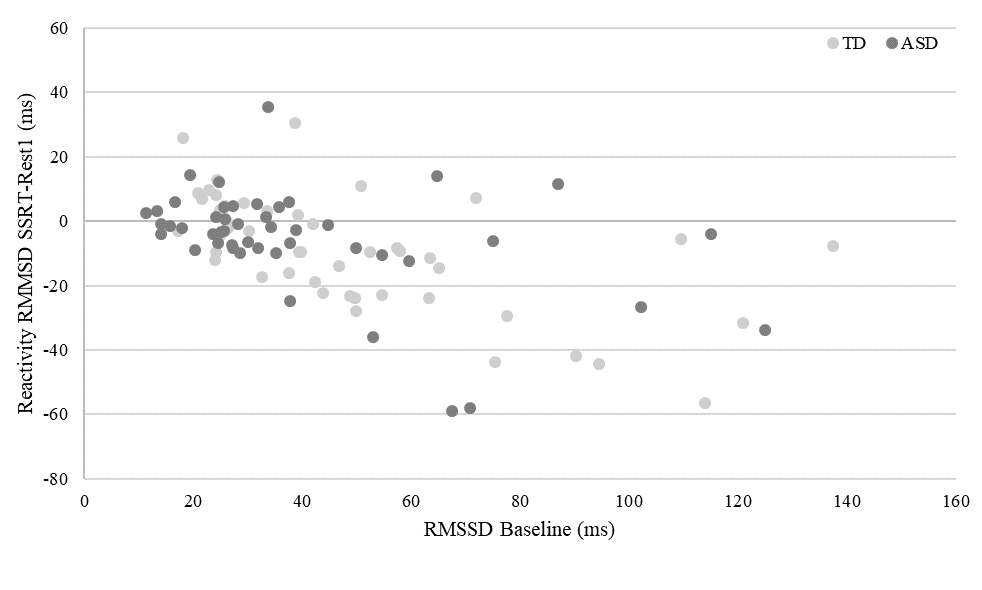

**Supplementary Material C** Association between baseline cardiac vagal modulation and cardiac vagal reactivity during the SSRT based on Spearman’s correlation analysis (ASD: *r_s_*=-.395; TD: *r_s_*=-.646; *p*<.01). Note: to ease the interpretability of this graph, raw baseline RMSSD values were used.

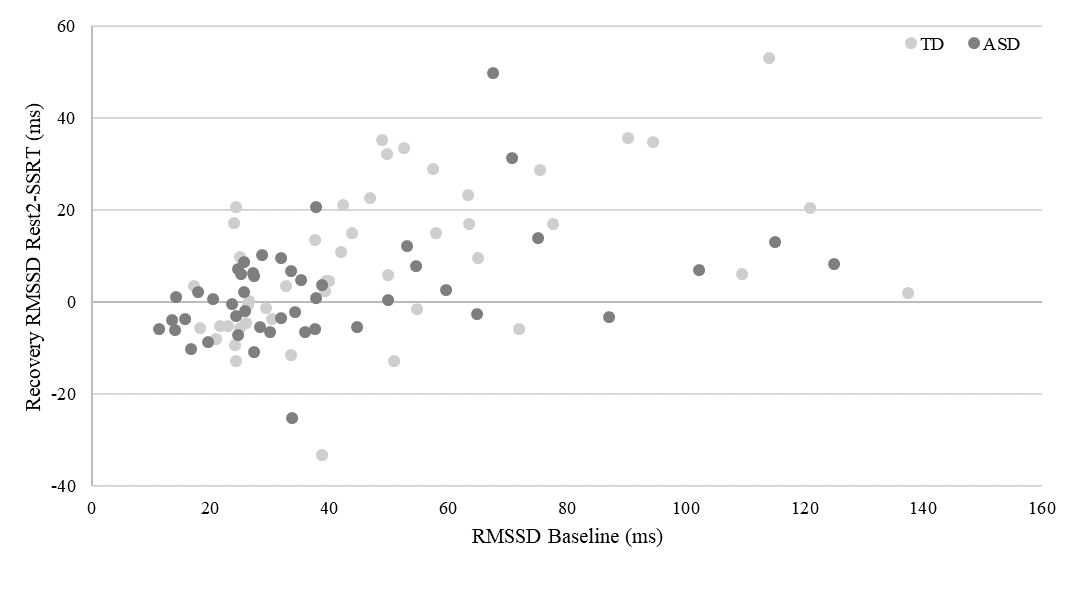

**Supplementary Material D** Association between baseline cardiac vagal modulation and cardiac vagal recovery following the SSRT based on Spearman’s correlation analysis (ASD: *r_s_*=.456; TD: *r_s_*=.530; *p*<.01). Note: to ease the interpretability of this graph, raw baseline RMSSD values were used
